# Gender-Based Analysis of PTSD in Addiction

**DOI:** 10.1101/2025.09.25.25336653

**Authors:** Imane Katir, A. Korchi, H. Zarouf, S. Belbachir, M. Sabir, A. Ouanass

## Abstract

**Background:** Post-Traumatic Stress Disorder (PTSD) affects 8-10% of trauma-exposed individuals globally (2), with significant gender disparities in prevalence and clinical presentation. The relationship between PTSD and substance use disorders creates complex comorbidity patterns requiring gender-sensitive approaches.

**Objective:** To examine gender-specific differences in PTSD symptom expression and addiction patterns among patients in addiction treatment settings.

**Methods:** Cross-sectional epidemiological study with retrospective components examining 30 participants (60% women, 40% men) aged 18 and above with confirmed PTSD diagnoses and comorbid addictive disorders. Standardized questionnaires including PCL-5 assessments were combined with systematic medical record analysis over eight months. Statistical analyses employed Mann-Whitney U tests, Chi-square tests, and logistic regression models.

**Results:** Women demonstrated higher PCL-5 total scores (56.3 vs 52.6) and greater intrusive symptoms (17.0 vs 14.8). Cannabis emerged as the primary self-medication substance (26.7% of sample), with 60% of users employing it for trauma-related symptoms. Women showed higher rates of cannabis use for trauma (70% of cannabis users), while men demonstrated greater stimulant use (70% of stimulant users). Psychiatric antecedents were prevalent (87%) with significant association to elevated PCL-5 scores (p < 0.05).

**Conclusions:** Distinct gender-specific patterns emerged in trauma response and addiction behaviors. Women demonstrated internalized symptom presentations with higher intrusive symptoms, while men showed externalized patterns with greater symptom variability. These findings support the need for gender-sensitive screening and integrated treatment approaches in addiction settings.

## Introduction

Post-Traumatic Stress Disorder (PTSD) represents a major public health challenge with significant global impact. According to the World Health Organization, approximately 70% of the world’s population reports exposure to at least one potentially traumatic event during their lifetime, with 8-10% subsequently developing PTSD (2,16,17). This complex psychiatric condition manifests through a constellation of symptoms including intrusive re-experiencing (flashbacks, nightmares), persistent avoidance of trauma-related stimuli, negative cognitive and emotional alterations, and marked neurovegetative hyperactivation (4,5).

Initially described as “traumatic neurosis” in war veterans, PTSD has evolved into a distinct diagnostic category within modern nosographic classifications, now featured in the DSM-5-TR under “Trauma and Stressor-Related Disorders” (4,5). The disorder’s heterogeneous nature reflects the complex interplay between traumatic exposure and individual vulnerability factors spanning biological, psychological, and environmental domains according to the biopsychosocial model (25,26).

Gender emerges as a critical determinant in both PTSD development and clinical presentation (6,7,8). Despite men’s statistically higher exposure to potentially traumatic events, women demonstrate approximately twice the risk of developing PTSD (8-12% versus 3-6%) (6,7,18). This gender disparity reflects multifaceted interactions including differences in trauma types experienced (sexual violence, domestic abuse being more prevalent in women), hormonal and neurobiological vulnerabilities, and psychosocial factors that fundamentally shape how psychological distress is perceived, expressed, and managed (7,8,18).

Societal gender norms significantly influence symptom expression and help-seeking behaviors (9,10,11). Women are generally encouraged to verbalize emotional distress, while men are often socialized toward emotional concealment, potentially contributing to underdiagnosis of PTSD in males when symptoms manifest through externalizing behaviors such as anger, violence, or risk-taking rather than explicit complaints (9,10,11). This gendered bias extends to clinical practice, where female presentations (anxiety, somatization) may be misinterpreted as mood disorders, while male behaviors (substance abuse, aggression, isolation) can mask underlying post-traumatic distress (9,10,11).

### PTSD and Addictive Disorders: Understanding the Clinical Interface

The relationship between PTSD and substance use disorders represents one of the most complex comorbidity patterns in psychiatric practice, with up to 50% of PTSD patients presenting concurrent addiction issues (12,13). This bidirectional association creates intricate clinical challenges where PTSD increases addiction risk while substance use can mask or exacerbate traumatic symptoms (12,13,14). In clinical addiction settings, 30-60% of patients present comorbid PTSD, with substance use frequently serving as emotional self-regulation according to the self-medication hypothesis (14,15).

Different substances serve distinct self-medication functions in PTSD management(13,14,58,59). Alcohol is predominantly used for its anxiolytic and sedative properties, particularly effective for hypervigilance and insomnia (13,58). Cannabis is frequently employed to manage anxiety and nightmares, though chronic use may paradoxically worsen certain symptoms. Stimulants like cocaine and amphetamines are utilized to counter the affective numbing and anhedonia characteristic of PTSD, while opioids provide psychological numbing but carry extremely high addiction potential (13,14,58,59).

### Gender-Differentiated Vulnerability and Expression

Gender emerges as a fundamental determinant in both PTSD development and its addictive comorbidities (6,7,8,52,53,54). Women demonstrate heightened biological vulnerability through hormonal influences on the hypothalamic-pituitary-adrenal axis and elevated inflammatory markers (IL-6) linked to violence-related trauma (7,8). They experience peak PTSD prevalence in their early fifties, with female-to-male ratios reaching 3:1 at certain ages (6,7).

The “quantity-quality paradox” defines gender differences in trauma exposure: while men face greater overall traumatic exposure, women develop PTSD twice as frequently due to experiencing predominantly interpersonal traumas (sexual violence, domestic abuse, childhood sexual abuse) that carry higher PTSD risk (6,7,20,21). Men typically encounter non-interpersonal traumas (accidents, natural disasters, physical assaults) and show peak prevalence in their early forties (6,7,20,21).

Symptom manifestation varies significantly by gender (6,7,8,52,53). Women typically present with internalized symptoms including frequent nightmares, intense flashbacks, marked avoidance behaviors, pronounced hypervigilance and anxiety, dissociative episodes, and co-occurring depressive and anxiety symptoms (6,7,18,52). Men more commonly display externalized presentations: underreported flashbacks, minimized avoidance, hypervigilance associated with anger and aggression, masked depressive symptoms through external behaviors, under-diagnosed anxiety, overt anger and impulsivity, substance use masking dissociation, and frequent risk-taking behaviors (9,10,11,52,53).

Understanding these gender-specific presentations becomes essential for improving identification, diagnosis, and therapeutic interventions in addiction treatment settings, where mixed symptomatology often remains unrecognized within standardized care protocols (77,79,80).

## Methods

### Study Design and Setting

This cross-sectional epidemiological study with retrospective components was conducted in an urban addiction treatment setting over an eight-month period. The research examined gender influences on PTSD clinical expression and addiction profiles among patients receiving addiction treatment services.

### Participants

The study targeted patients aged 18 and above with confirmed PTSD diagnoses and comorbid addictive disorders. A total of 30 participants were included in the final analysis, comprising 18 women (60%) and 12 men (40%) with a median age of 34 years.

### Data Collection

The methodology combined standardized questionnaires with systematic medical record analysis. Prospective consultation assessments were conducted alongside retrospective archival data analysis to capture comprehensive clinical profiles.

### Instruments

Primary assessment tools included:

- **PCL-5 (PTSD Checklist for DSM-5):** Standardized assessment for PTSD symptom severity across four symptom clusters (re-experiencing, avoidance, negative cognitive/mood alterations, hyperarousal)
- **Structured clinical interviews:** For demographic data, substance use patterns, and trauma history
- **Medical record analysis:** For psychiatric history, medication use, and treatment outcomes

### Statistical Analysis

Statistical analyses employed:

- **Mann-Whitney U tests:** For symptom score comparisons between gender groups
- **Chi-square tests:** For categorical associations
- **Logistic regression models:** To examine relationships between gender, PTSD severity, and specific addiction patterns

Statistical significance was set at p < 0.05. Data analysis addressed the temporal relationship between PTSD and addiction development, with approximately 60-75% of cases following a pattern where PTSD precedes addiction development (12,13).

### Ethical Considerations

The study protocol adhered to ethical guidelines for clinical research, with appropriate institutional review and participant consent procedures.

## Results

### Sample Demographics and Characteristics

The study examined 30 participants with a slight female predominance (60% women, 40% men) and a median age of 34 years. Despite high educational attainment (76.7% university graduates), unemployment affected 23.3% of participants, with striking gender disparities: 33.3% of men were unemployed versus only 16.7% of women. Men were predominantly single (83.3%), while women showed greater relationship diversity (38.9% single, 33.3% married, 27.8% in relationships).

### Medical and Psychiatric History

Women demonstrated significantly higher rates of medical-surgical history (77.8% vs 16.7% in men), while psychiatric antecedents were prevalent across both groups (72.2% women, 58.3% men) (22,23). Current psychotropic medication use was common (60% overall), with higher rates among women (66.7% vs 50%). Family psychiatric history was remarkably high across both genders (70% overall), suggesting significant genetic vulnerability in this population (25,41).

### Addiction Patterns and Self-Medication Behaviors

Cannabis emerged as the primary self-medication substance, affecting 26.7% of the sample, with 60% of cannabis users employing it specifically for trauma-related symptoms (14,59).

Women showed higher rates of cannabis use for trauma (70% of cannabis users were female), while men demonstrated greater use of stimulants (70% of stimulant users were male).

Alcohol followed as the second most common trauma-related substance (20% of sample), though primarily used occasionally (66.7% of trauma-related cases), suggesting situational rather than systematic self-medication (13,58). Tobacco/nicotine showed daily consumption patterns (66.7% of trauma cases), indicating established dependence as continuous emotional regulation.Behavioral addictions (gambling, internet use) demonstrated exclusively daily patterns (80%) when trauma-related, suggesting their use as avoidance and dissociation mechanisms against intrusive memories.

### PTSD Symptom Expression by Gender

PCL-5 assessment revealed nuanced gender differences in symptom presentation:

### Re-experiencing/Intrusion Symptoms (Cluster B)

Women scored higher (17.0 vs 14.8), with both groups in the “moderate to severe” range (6,7). Women reported more frequent nightmares, intense flashbacks, and invasive memories.

### Avoidance Symptoms (Cluster C)

Scores were comparable between genders (8.35 women,8.23 men), both indicating moderate avoidance behaviors with low variability, suggesting uniform responses to trauma reminders.

### Cognitive and Mood Alterations (Cluster D)

Women showed slightly higher scores (17.9 vs 16.5), but men demonstrated greater variability (SD 8.29 vs 6.59), indicating more heterogeneous symptom profiles in areas like negative beliefs, emotional numbing, and guilt (52,53).

### Hyperarousal and Reactivity (Cluster E)

Scores were nearly identical (13.1 women, 13.2 men), but men showed greater variability (SD 6.26 vs 3.54), suggesting more disparate experiences in hypervigilance, irritability, and sleep disturbances (52,53).

### Total PCL-5 Scores

Women scored higher overall (56.3 vs 52.6), with both groups exceeding clinical significance thresholds (≥33) (1,3). However, confidence intervals overlapped substantially, indicating the difference may be clinically meaningful but not statistically significant.

### Substance-Specific Usage Patterns

#### High Trauma-Related Self-Medication Substances

- Cannabis: 26.67% trauma-linked usage (highest rate), with 60% of cannabis users employing it specifically for trauma symptoms (14,59)
- Alcohol: 20% PTSD self-medication, predominantly occasional use suggesting situational coping (13,58)
- Tobacco: 14.28% trauma-related consumption, but 39.29% severe addiction indicating continuous emotional regulation

#### Low Trauma-Involvement Substances

- Stimulants: Minimal usage (10.34%) with virtually no trauma-related self-medication (3.45%)
- Opioids: 10% severe addiction but 0% trauma self-medication
- Diverted medications: 6.67% trauma usage with iatrogenic risk concerns

#### Emerging Behavioral Addictions

- Compulsive shopping: Notable prevalence (23.33%) with 3.33% trauma-linked, predominantly affecting women
- Internet/social media: 20% problematic usage, 6.67% trauma self-medication
- Pathological gambling: Low prevalence (10%) but 6.67% trauma avoidance mechanism
- Video gaming: Recreational use only (10%)

## Discussion

### Sociodemographic Profile and Clinical Context

The study reveals a highly educated urban population (76.7% university graduates) with a median age of 34 years, demonstrating that PTSD affects all socioeconomic strata (31,76). However, significant gender disparities emerge: men show higher unemployment rates (33.3% vs 16.7%) despite similar education levels and are predominantly single (83.3% vs 38.9% for women). These differences suggest gendered impacts of PTSD on social and professional functioning, potentially reflecting differential coping strategies and social support utilization (9,10,11).

### PTSD Symptom Expression and Severity

PCL-5 scores were substantially elevated across both groups (52.8 for men, 59.0 for women), far exceeding the diagnostic threshold of 33 points, indicating severe symptomatology consistent with addiction treatment populations (1,3). Women demonstrated higher intrusive symptom scores (PCL-4, PCL-5) and hyperarousal symptoms (PCL-17), supporting previous research on gender differences in PTSD intensity (6,7,52). The non-normal distribution of male scores suggests greater symptom heterogeneity, potentially reflecting diverse coping strategies or underreporting of certain symptoms (9,10,11,52,53).

### Psychiatric Comorbidity and Clinical Complexity

The extremely high prevalence of psychiatric antecedents (87%) confirms the clinical complexity of this population (22,23,33,78). The significant association between psychiatric history and elevated PCL-5 scores (p < 0.05) demonstrates cumulative effects of mental health vulnerabilities (25,41). This finding aligns with neurobiological models suggesting preexisting vulnerabilities may increase both PTSD risk and severity of associated disorders (26,27,47,71).

### Self-Medication Hypothesis and Substance Use Patterns

The findings strongly support the self-medication hypothesis, particularly for cannabis use (14,59). Cannabis emerged as the central PTSD self-medication strategy requiring specific clinical attention, while tobacco represented the highest severe addiction risk. The emergence of behavioral addictions as new avoidance modalities, particularly among women, necessitates enhanced clinical vigilance.

### Gender-Specific Clinical Profiles

#### Female Pattern

- More intense, stable symptom presentation with predominant re-experiencing (PCL-B) and mood alterations (PCL-D) (6,7,52)
- Alcohol and cannabis self-medication (20% and 26.67% respectively) (13,14,58,59)
- Greater reliance on behavioral avoidance strategies (compulsive shopping, problematic social media use)
- Earlier trauma exposure (average age 19.5 vs 22.1 for men), predominantly interpersonal traumas (6,7,18)

#### Male Pattern

- More heterogeneous symptoms, particularly in hyperarousal clusters (PCL-E) (52,53)
- Less trauma-specific self-medication but higher rates of dependence addictions (tobacco, opioids)
- Externalized symptom expression with dominant physical dependencies (9,10,11)
- Greater cannabis consumption overall (69% vs 29% for women)

### Neurobiological Implications

Gender differences appear linked to distinct cerebral activation patterns, particularly involving amygdala and prefrontal cortex circuits (26,27,47,71). Women utilize cannabis and alcohol primarily for intrusive symptom regulation, while men demonstrate addictions less directly trauma-related but more oriented toward stimulation-seeking or behavioral compensation. These patterns suggest differentiated neurobiological mechanisms in PTSD self-medication according to substance type (26,27,47,71).

### Limitations

The study’s limitations include the relatively small sample size (n=30), single-center design, and potential selection bias inherent in addiction treatment settings. The cross-sectional design limits causal inferences, and the high educational level of participants may not be representative of broader PTSD populations (31,76).

### Clinical Implications and Treatment Considerations

The findings support implementing gender-sensitive, individualized therapeutic approaches that account for distinct symptom presentations, self-medication patterns, and substance preferences (77,79,80,91,99,107,115,123,131,139,147). The identified dual vulnerability(addiction + PTSD) affects significant patient proportions (16.67% for cannabis, 6.67% for alcohol), reinforcing needs for early screening and integrated treatment approaches (24,29,46,69,74,86,95,103,106,111,119,122,127,135,138,143,146).

## Conclusions

This study reveals distinct gender-specific patterns in PTSD symptom expression and addiction behaviors within addiction treatment settings. Women demonstrated more internalized symptom presentations with higher intrusive symptoms and greater use of cannabis for anxiety management, while men showed more externalized patterns with greater symptom variability, higher stimulant use, and potential underreporting of emotional distress (6,7,52,53).

The results advocate for developing personalized, gender-specific, and transdiagnostic therapeutic protocols combining psychotraumatological, addiction, and psychosocial interventions (77,79,80,91,99,107,115,123,131,139,147). Cannabis emerges as a central self-medication strategy requiring specific clinical attention (14,59), while the emergence of behavioral addictions, particularly among women, necessitates enhanced clinical vigilance.

These findings establish foundations for future research exploring individual trajectories, underlying neurobiological mechanisms, and comparative efficacy of targeted intervention strategies for these differentiated clinical profiles (26,27,47,71,95,103,111,119,127,135,143).

The high prevalence of family psychiatric history (70%) underscores the importance of considering genetic vulnerability in treatment planning (25,41), while the complex interplay between PTSD and addiction requires integrated, gender-sensitive approaches to optimize therapeutic outcomes (24,29,46,69,74,86,106,122,138,146).

## Data Availability

All data produced in the present work are contained in the manuscript.

